# Effects of Equine-Assisted Therapy on Recovery after Stroke – A Systematic Review

**DOI:** 10.1101/2023.05.09.23289710

**Authors:** Bettina Hanna Trunk, Alireza Gharabaghi

## Abstract

**Background:** Equine-Assisted Therapy (EAT) can boost well-being and recovery of patients with neurological or psychiatric disorders.

**Objective:** The goal of this systematic review is to gain a better understanding of the effects of EAT on recovery after stroke.

**Methods:** A systematic literature search was performed in the following databases: PubMed, Web of Science and Scopus. Furthermore, reference lists from the articles included were screened. English-written articles published between 2000-2023 that reported on health-related effects of EAT (applied with both horses and riding simulators) on stroke recovery in patients aged between 18 and 85 were included. Methodological quality was assessed by the Mixed Methods Appraisal Tool.

**Results:** Following the screening of 2030 and retrieval of 33 articles respectively, 14 studies were included in this systematic review (437 patients, mean age range: 40 – 70 years). Since several of these studies lacked important methodological information, the overall methodological quality varied. Thirteen studies reported physical findings (balance, gait, postural coordination, activities of daily living, lower extremity motor impairment, motor function and hand strength), and seven studies reported further health-related outcomes (cognition, quality of life, depression and perception of the intervention, muscle thickness and trunk muscle activity). The findings suggest positive effects of EAT on stroke recovery in different health-related outcomes, whereas the most consistent beneficial effects were reported for balance and gait.

**Conclusion:** EAT appears to be a promising multimodal intervention for the recovery of different functions after stroke. However, evidence is sparse and methodological quality limited. Future research should investigate the effects of EAT on stroke recovery more systematically.

## Introduction

One out of four people over the age of 25 are expected to suffer from a stroke within their lifetime. (Feigin *et al*., 2022) Stroke is a devastating disease which can lead to various, severe, and persistent symptoms, such as hemiparesis of the upper and lower extremity, spasticity, aphasia, and other cognitive impairments. (Brewer *et al*., 2013) Due to natural reorganization processes, patients can, to some extent, recover lost functions within the first six months following a stroke. However, after this period, no further recovery is expected with current therapies. (Murphy and Corbett, 2009; Grefkes and Ward, 2014) Persistent disabilities have a drastic impact on the patients’ quality of life, as they are often dependent on caregivers, even for simple activities of daily life. (Kim *et al*., 1999; Feigin *et al*., 2015) Thus, novel interventions are required. (Achten *et al*., 2012)

Multimodal interventions, such as Equine-Assisted Therapy (EAT), have been proposed to engage patients in concurrent physical, sensory, cognitive and social activities, therefore supporting recovery from multiple symptoms. (Pekna *et al*., 2012, Bunketorp-Käll *et al*., 2017*b*) While riding a horse at walking speed, the three-dimensional movement of the horse’s back leads to a rhythmic sensorimotor stimulation of the patients that is comparable to human gait. (Uchiyama *et al*., 2011; Garner and Rigby, 2015, Bunketorp-Käll *et al*., 2017*b*; Guindos-Sanchez *et al*., 2020) At around 70-100 steps/minute, the rider is passively moved. (Uchiyama *et al*., 2011) Patients must therefore constantly adjust to small postural changes. (Rigby and Grandjean, 2016) EAT can be performed with both horses and riding simulators. (Baek and Kim, 2014, Bunketorp-Käll *et al*., 2017*b*) The former in particular provides an enriched environment, ultimately leading to a higher training motivation and improved psychological outcome. (Bunketorp-Käll *et al*., 2017*b*; Vive *et al*., 2022; Ward *et al*., 2022)

While the exact mechanisms of action have not been understood yet, several theories have been suggested in this regard. (Lewis, 2020) For example, the similarity of sensorimotor stimulation during EAT and human gait has been suggested to strengthen the muscles of the rider, ultimately leading to improvements in gait. (Beinotti *et al*., 2013; Lewis, 2020) Furthermore, the rider needs to continuously adjust to the horses’ movement (McGibbon *et al*., 2009; Uchiyama *et al*., 2011; Garner and Rigby, 2015, Bunketorp-Käll *et al*., 2017*b*; Guindos-Sanchez *et al*., 2020) which may train their vestibular system and postural control, ultimately improving their balance. (de Mello *et al*., 2022) Moreover, the horses’ girth may lead to a stretch of adductor muscles of hips and legs and decrease spasticity. (McGibbon *et al*., 2009) Furthermore, horses have a higher body temperature than humans which may also lead to decreased spasticity and hypertonicity. (Lewis, 2020)

Benefits of EAT following severe neurological disability or illness have previously been reported (Viruega *et al*., 2022); usually in children with cerebral palsy (Deutz *et al*., 2018; Guindos-Sanchez *et al*., 2020; Heussen and Häusler, 2022), but also in patients with multiple sclerosis (Stergiou *et al*., 2017), spinal cord injury (Lechner *et al*., 2007), ADHD (Hyun *et al*., 2016; Yoo *et al*., 2016), chronic pain (Collado-Mateo *et al*., 2020), Down syndrome (Portaro *et al*., 2020), cancer (Viruega *et al*., 2023), or stroke (Beinotti *et al*., 2013, Bunketorp-Käll *et al*., 2017*b*). EAT was found to benefit physical function, i.e., improved gait, balance, or gross motor function, as well as to decrease spasticity. (Guindos-Sanchez *et al*., 2020; Badin *et al*., 2022) In addition, psychological factors, such as quality of life and depression were found to be improved in various cohorts. (Badin *et al*., 2022; Viruega *et al*., 2023)

Consequently, EAT appears to be a promising multimodal intervention that may facilitate recovery of multiple symptoms in stroke patients. (Bunketorp-Käll *et al*., 2017*b*) In addition, EAT provides a much higher training intensity than other interventions. This has been proposed to be vital for boosting further recovery in chronic stroke patients (Krakauer *et al*., 2012; Lohse *et al*., 2014; Solomonow-Avnon and Mawase, 2019; Ward *et al*., 2019). However, previous research of EAT in stroke patients is sparse and, to the best of our knowledge, no systematic review has yet synthesized the effects of EAT on different domains of stroke recovery yet. Moreover, it remains unclear whether the effects differ when EAT is applied with horses or riding simulators.

We therefore aim to 1) provide an overview of the existing literature on EAT in stroke patients; 2) evaluate the methodological quality of these studies; 3) synthesize the effects of EAT on health-related outcomes during recovery after stroke and; 4) assess the differences in outcome when applying EAT with horses or riding simulators.

## Methods

This systematic review was conducted in accordance with the Physiotherapy Evidence Database (PRISMA) guidelines (Page *et al*., 2021) as well as the guidelines for synthesis without meta-analysis (SWiM) (Campbell *et al*., 2020). This review was not registered. Data search, extraction, synthesis, and quality assessment were done by one reviewer (BHT).

### Eligibility criteria

Inclusion criteria were defined based on the PICO approach (population, intervention, control, outcome).

The patient population of interest were stroke patients between 18 and 80 years of age. We included intervention studies published between 2000 and 2023 in which EAT had been applied (independent of the precise intervention) in both horses and riding simulators. Furthermore, studies that included different patient cohorts were taken into consideration only if they were mainly comprised of stroke patients. Moreover, we took only those studies into account in which health-related outcomes (i.e., in physical, psychological, and physiological domains) were investigated. We did not consider manuscripts in any language other than English, conference work, theses, case reports, and study protocols.

### Search strategy

A search between January 1, 2000, and May 2, 2023 was conducted in the PubMed, Web of Science and Scopus databases. Furthermore, reference lists were cross-checked for potentially missed studies. Our search included search terms related to EAT and stroke and were connected using Boolean operators. The complete syntax for each library can be found in Supplemental Material.

### Selection process

All search results were downloaded to Mendeley with title, authors and abstract. Duplicates were then removed. Titles and abstracts were screened for eligibility. Full-text copies of the relevant studies were then downloaded and screened for eligibility.

### Data extraction

The following data were extracted from the studies selected: publication characteristics (authors, year, and location), aim of the study, study characteristics (age, sample size, time since stroke), intervention (details of experimental and control group, and dose of treatment), outcome measures, general findings. The data was organized in an Excel spreadsheet and labelled according to the health domain of interest.

### Data Synthesis

Results were then qualitatively synthesized. For synthesis, data was sorted regarding the study design (RCT and NRT) and then sorted regarding studies that applied the intervention in horses and in simulators. We furthermore investigated and synthesized results of physical and other health-related effects. If there was sufficient data for specific outcomes, harvest plots were created in order to assess heterogeneity of the data and to synthesize findings.

Within- and between group effects were furthermore described. For this, Cohen’s d effect sizes and respective 95% Confidence Intervals (CI) were computed (see Table 1). For within-group effects, i.e., changes over time (pre-to post-intervention), the timepoints immediately before and after the intervention or the nearest post timepoint were chosen. To account for differences in the direction of the scales, mean differences were multiplied by -1 for scales in which a higher number represents a better outcome. Therefore, positive effect sizes describe either favorable within-effects over time or between-effects for the experimental group (EG) compared to the control group (CG). Effects sizes (ES) of d = 0.10 – 0.30 were interpreted as small, of d = 0.30 – 0.50 as moderate, and of greater than 0.5 as large (Goulet-Pelletier and Cousineau, 2020).

### Quality assessment

The quality of the included studies was assessed using the Mixed Methods Appraisal Tool (MMAT) version 2018. (Hong *et al*., 2018) This tool was designed for evaluating the quality of studies that used mixed methods (randomized controlled trials, non-randomized trials, quantitative descriptive studies, and mixed methods studies) to be included in systematic reviews based on five quality criteria. (Hong *et al*., 2018) Criteria that were met were scored by a “Y”, whereas those that were not met were scored with a “N” or a “U” in the event of missing information. No studies were excluded based on their methodological quality.

## Results

### Study selection

Our systematic literature yielded 3107 articles. Then, 1077 duplicates were removed. After screening, 1996 articles were excluded, and 33 articles were retrieved and checked for eligibility. 17 articles met the inclusion criteria. As four reports reported outcomes from the same study (Bunketorp-Käll *et al*., 2017*b*, 2019, 2020), we included 14 studies in this review (Fig. 1). The studies involved are summarized in Tab. 1.

**Fig. 1.**
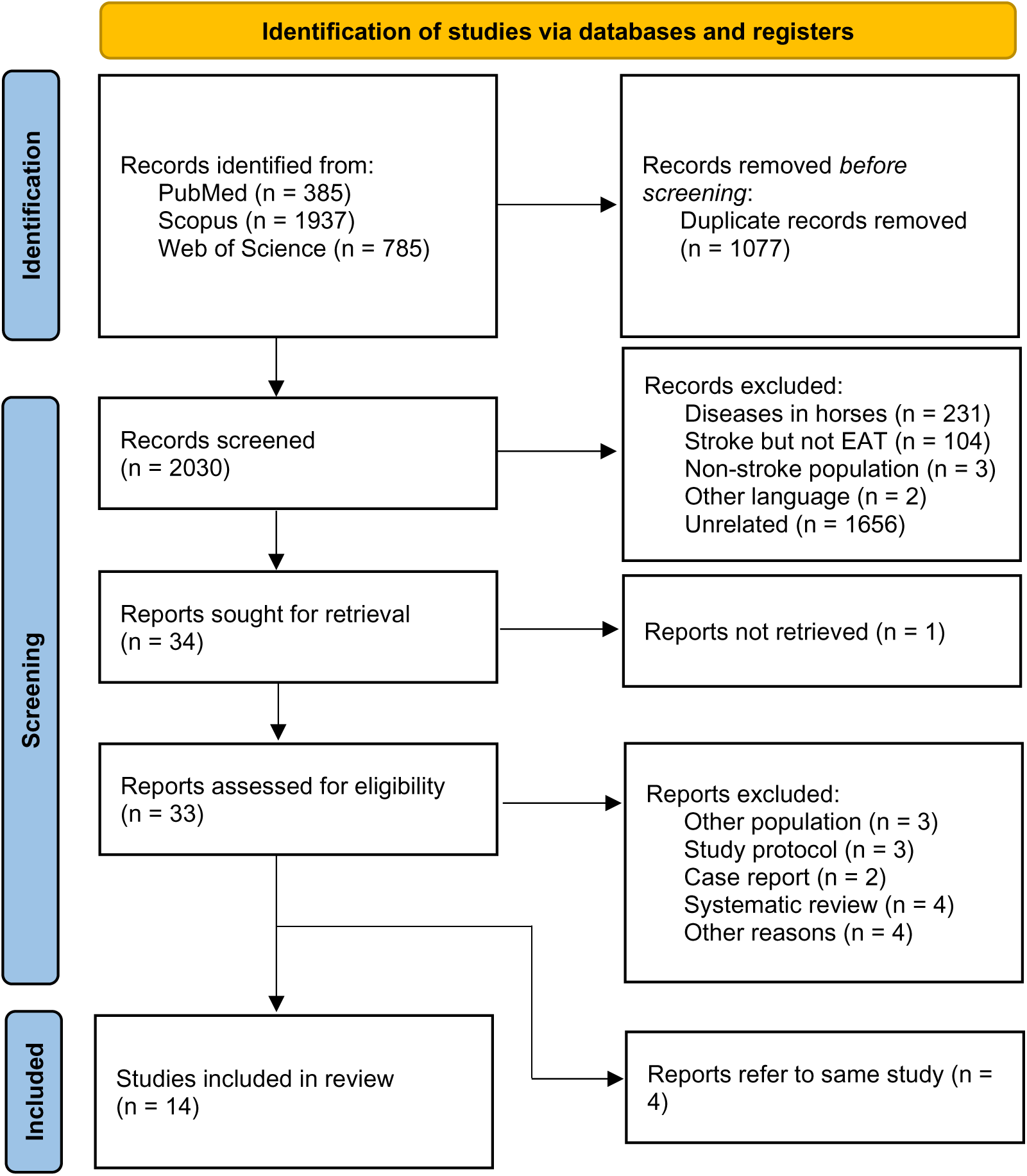
Flow diagram of enrolment based on PRISMA guidelines.

**Tab. 1.**
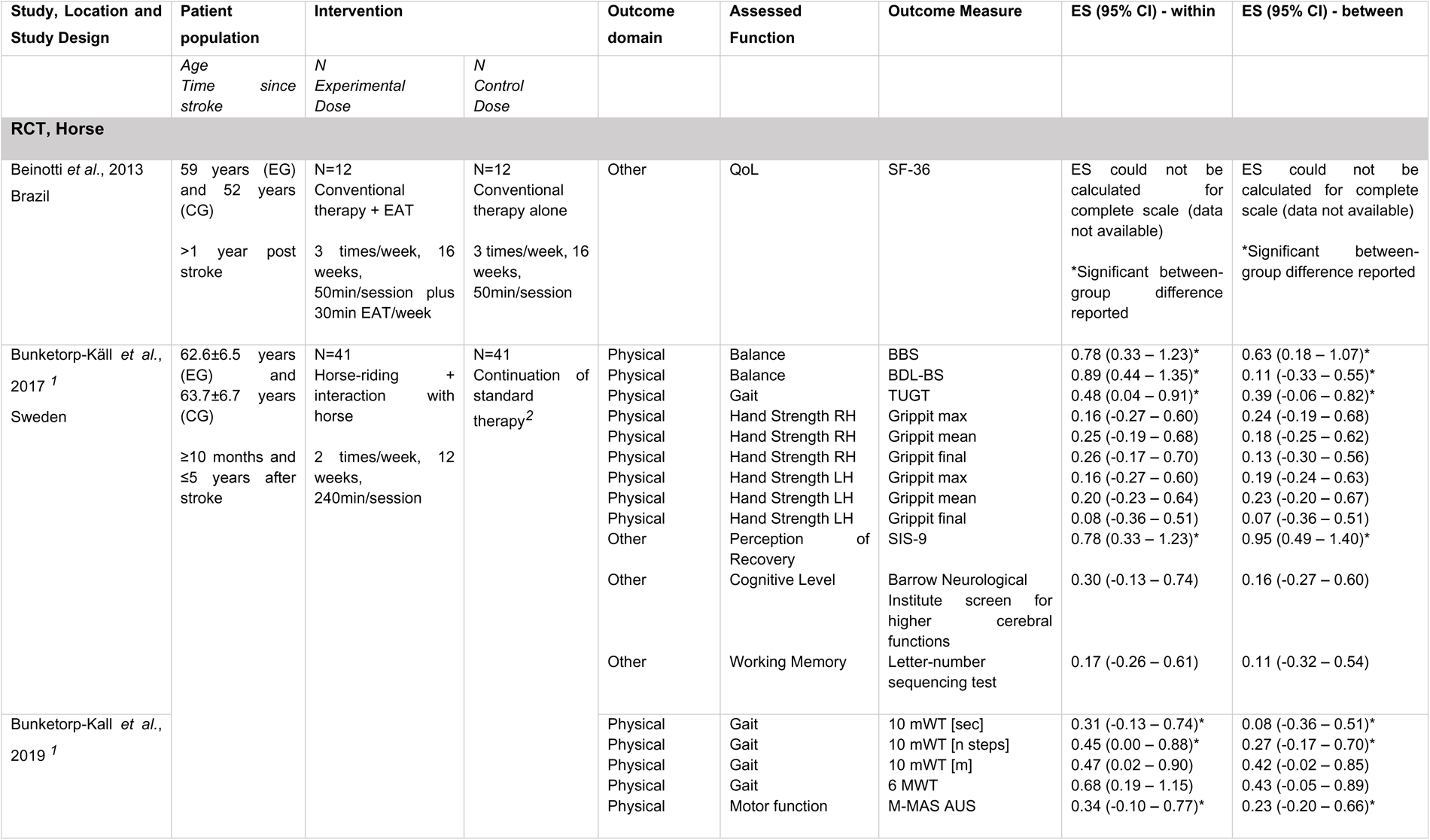
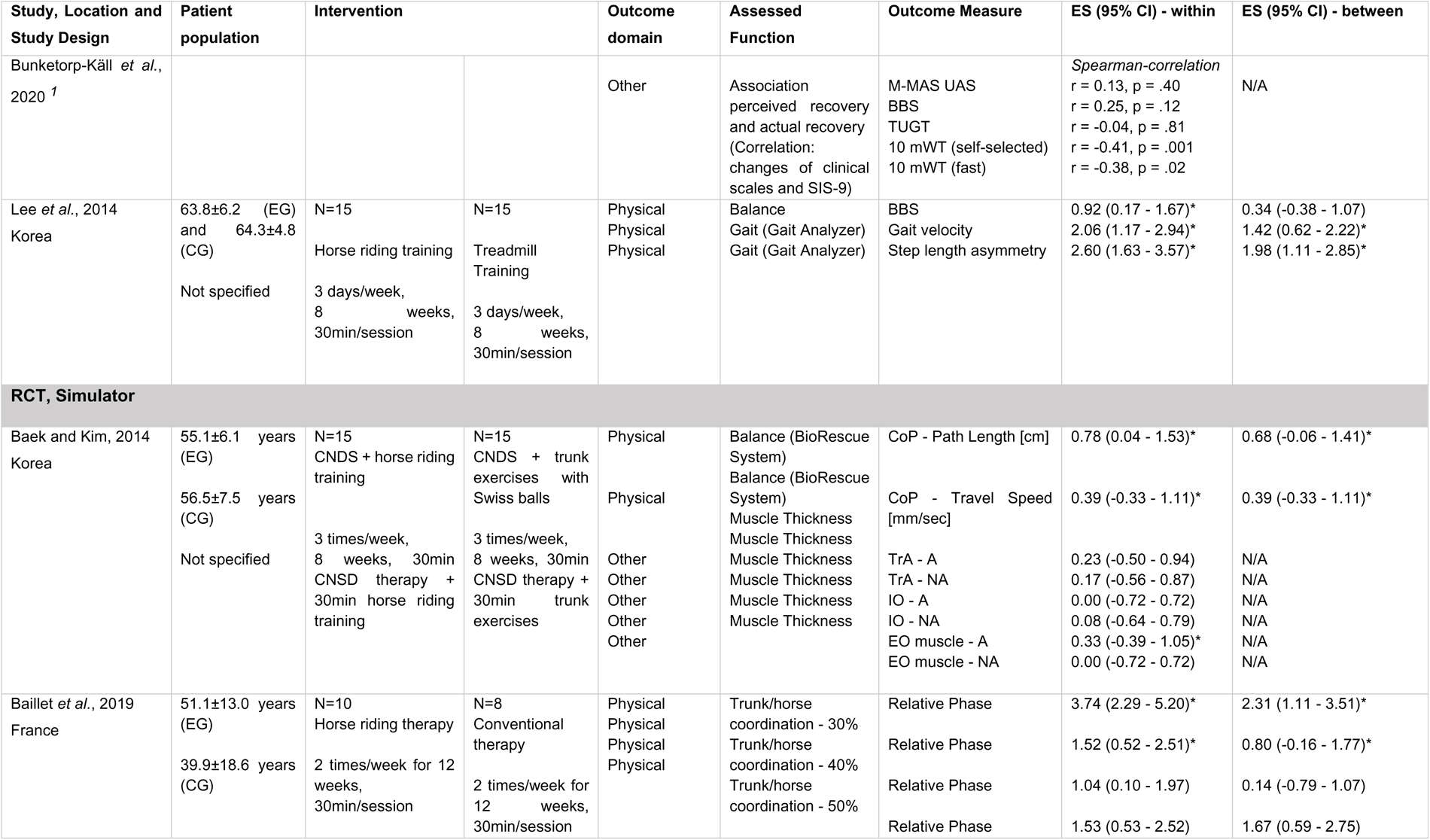
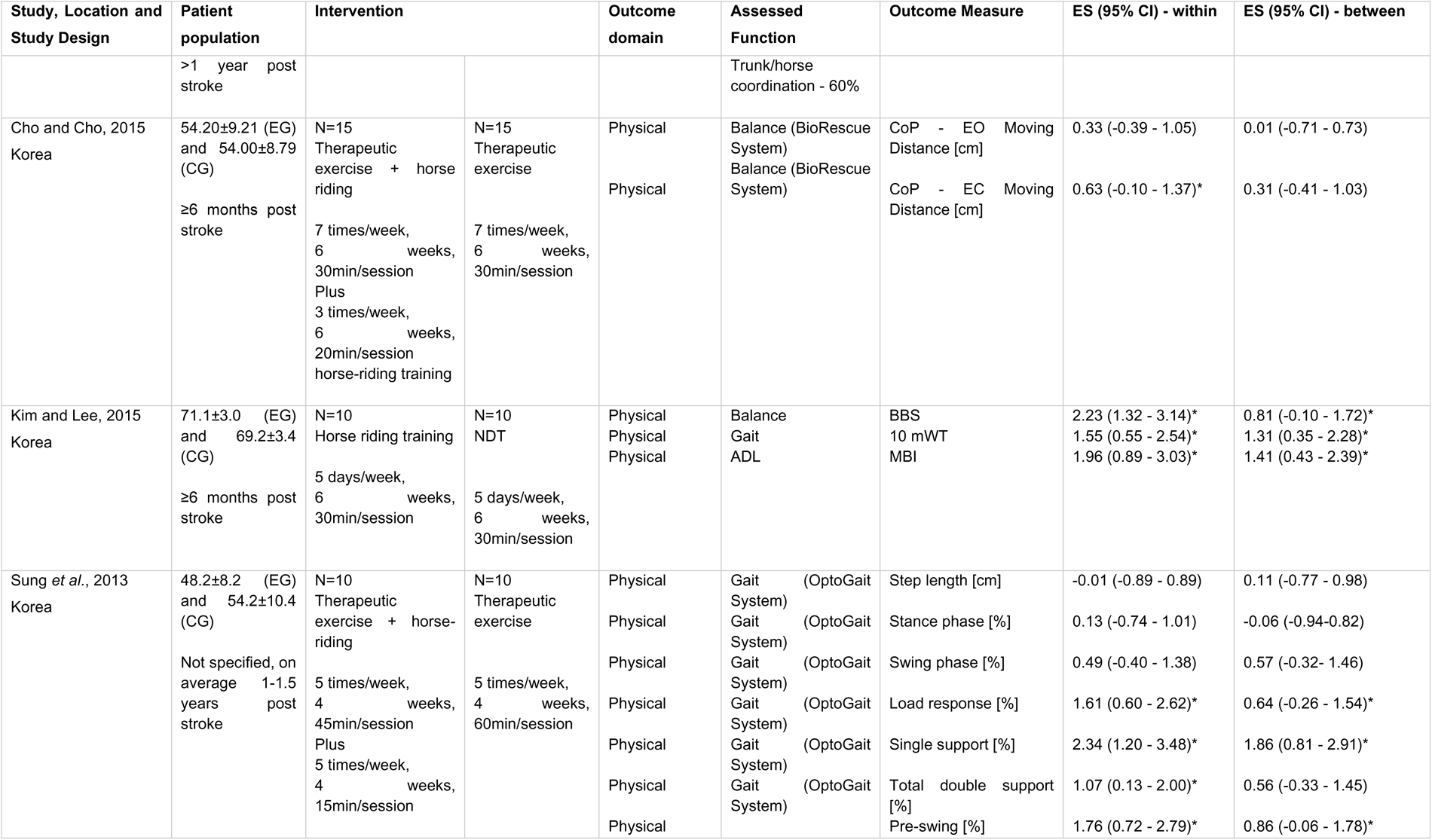
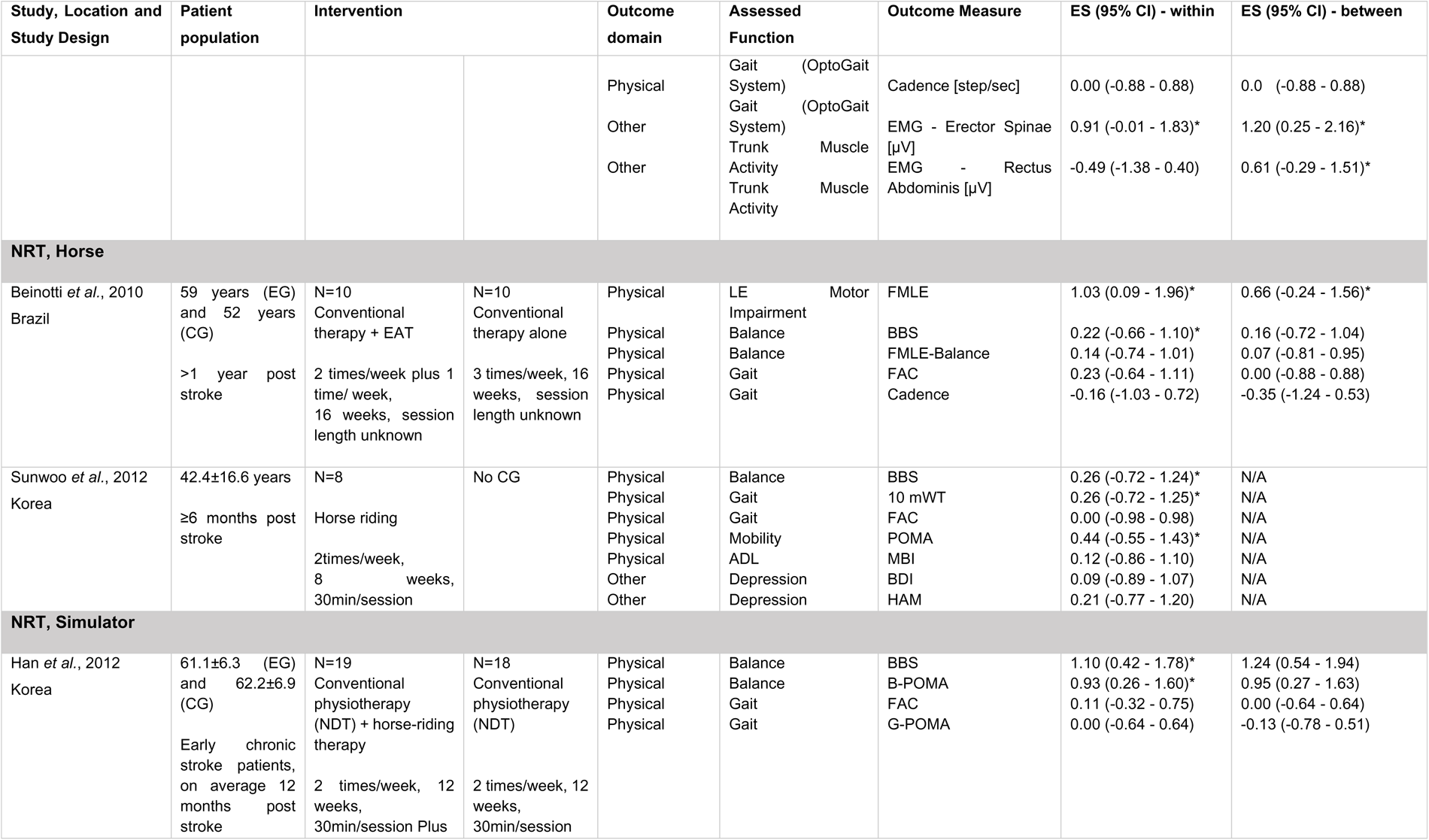
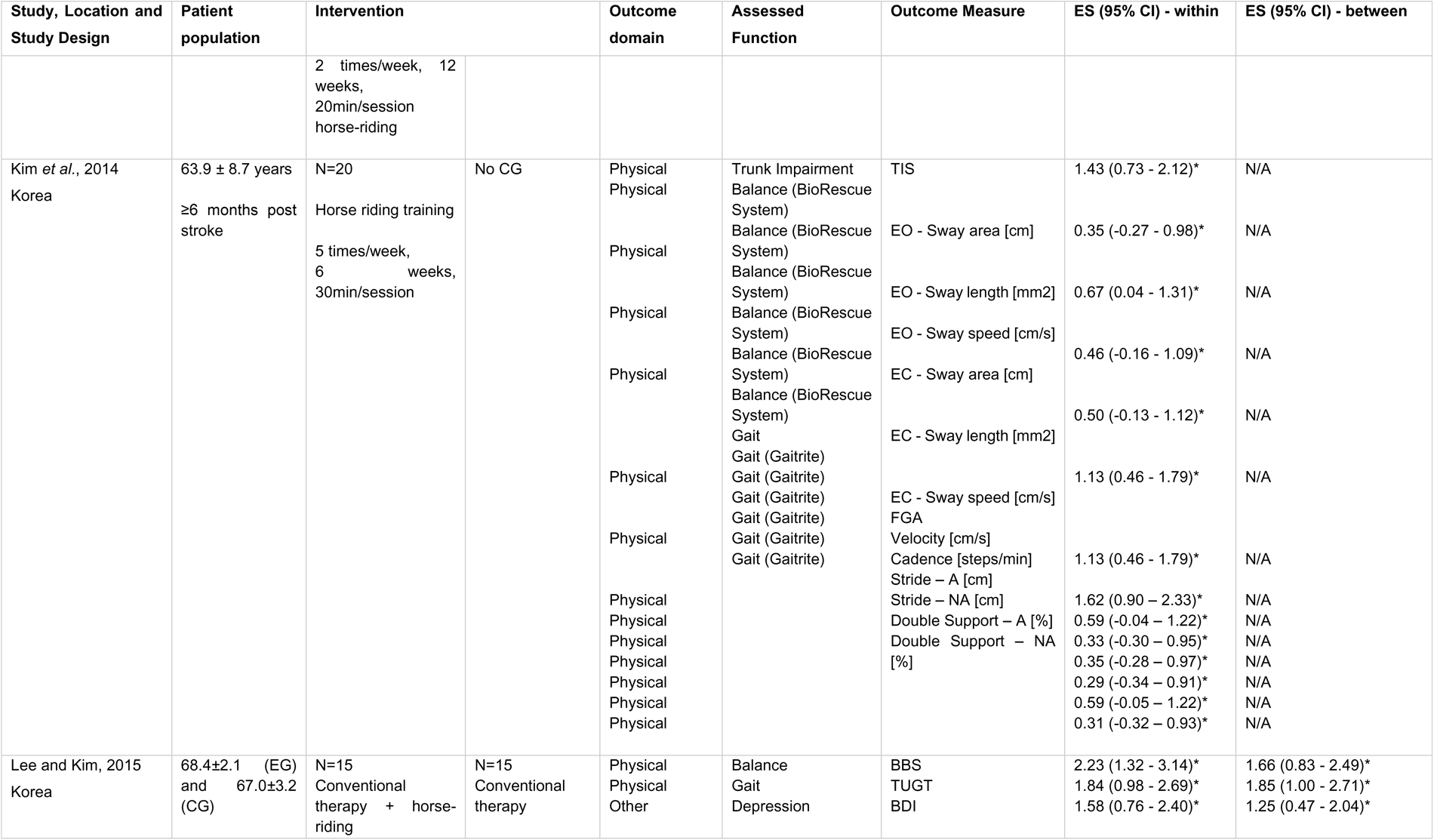
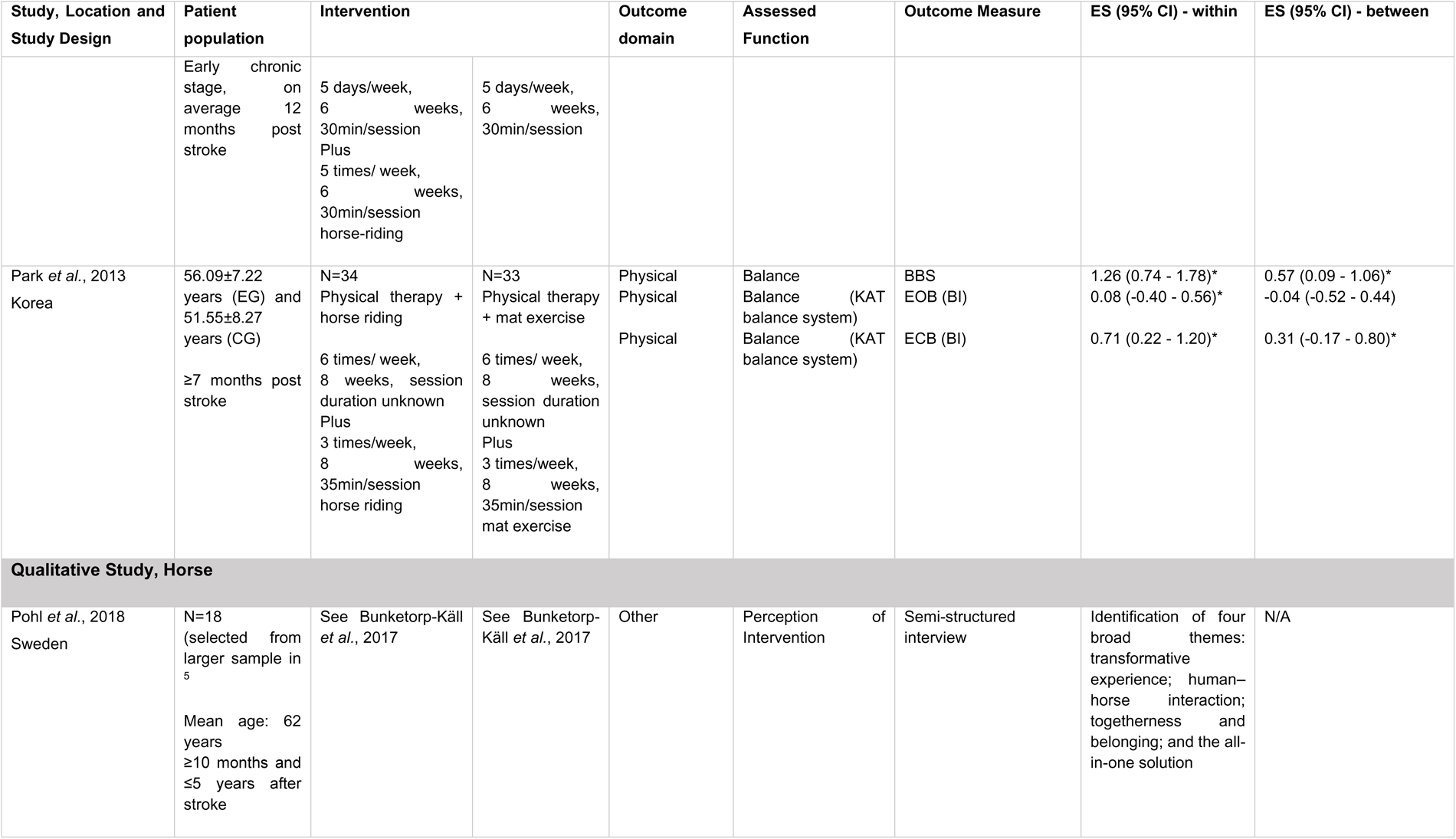
Overview of the studies included. This overview was sorted regarding the study design (RCT, NRT and Qualitative Study) and whether the intervention was applied with a horse or riding simulator. For each study, outcomes are sorted regarding the health-domain, i.e., physical, and other health-related outcomes. ^1^Findings from data of one study. ^2^Study included a second experimental condition. *Significant effect reported in paper. Abbreviations: A = Affected, ADL = Activities of daily living, BBS = Berg Balance Scale, BDL-BS = Bäckstrand, Dahlberg and Liljenäs Balance Scale, BDI = Becks Depression Inventory, BDL-BS = Bäckstrand, Dahlberg and Liljenäs Balance Scale, B-POMA = Balance Part of Performance Oriented Mobility Assessment, CG = control group, CNSD = Central Nervous System Developmental, COP = Center of Pressure, DRP = Discrete relative phases, EAT = Equine-Assisted Therapy, EC = eyes closed, EG = experimental group, EMG = Electromyography, EO = eyes open, EO muscle = External Oblique muscle, FAC = Functional Ambulation Category Scale, FGA = Functional Gait Assessment, FMLE = Fugl-Meyer Lower Extremity Scale, G-POMA = Gait part of Performance Oriented Mobility Assessment, HAM = Hamilton Depression Rating Scale, KAT = Kinesthetic Ability Trainer, IO = Internal Oblique, LE = Lower Extremity, LH = Left Hand, MBI = Modified Barthel Index, M-MAS = Modified Motor Assessment Scale according to Uppsala University Hospital, NA = Non-Affected, NRT = Non-randomized Trial, POMA = Performance-Oriented Mobility Assessment, QoL = Quality of Life, RCT = Randomized Controlled, RH = Right Hand, SF-36 = Medical Outcomes Study 36-item Short-Form health survey, SIS-9 = Stroke Impact Scale – Item 9, TIS = Trunk Impairment Scale, TrA = Transversus Abdominis, TUGT = Timed Up and Go Test, 10 mWT = Timed 10-meter walk test, 6 MWT = The six-minute walk test

### Subject characteristics

In total, data from 437 patients (mean age range: 40 – 70 years) were included in this systematic review. Sample sizes ranged from 8 to 123 patients. With the exception of two studies (Baek and Kim, 2014, Lee *et al*., 2014*a*), the stroke stage was specified. In one study, the minimum time since stroke was not reported, but the patients had suffered a stroke on average about one year earlier. (Sung *et al*., 2013) All other studies included patients at the chronic stage of stroke (i.e., > 6 months post stroke). In two studies (Sunwoo *et al*., 2012; Baillet *et al*., 2019), the majority of patients were stroke patients, although, a small number of traumatic brain injury and cerebral palsy patients were also included. All other studies involved stroke patients only.

### Study characteristics and methodological quality

The included studies were conducted in Brazil (Beinotti *et al*., 2010, 2013), France (Baillet *et al*., 2019), Korea (Han *et al*., 2012; Sunwoo *et al*., 2012; Park *et al*., 2013; Sung *et al*., 2013, Kim *et al*., 2014*a*, Lee *et al*., 2014*a*; Cho and Cho, 2015; Kim and Lee, 2015; Lee and Kim, 2015), and Sweden (Bunketorp-Käll *et al*., 2017*b*; Pohl *et al*., 2018; Bunketorp-Käll *et al*., 2019, 2020).

Study design and methodological quality, assessed by MMAT, are displayed in Supplemental Material (Tab. 2). Data from eight Randomized Controlled Trial (RCT), six Non-Randomized Trial (NRT). Moreover, one report referring to data of one RCT reported qualitative outcome and was therefore assessed separately. The overall study quality varied between the studies. Specifically, seven reported studies (Sunwoo *et al*., 2012; Beinotti *et al*., 2013, Kim *et al*., 2014*a*, Bunketorp-Käll *et al*., 2017*b*; Pohl *et al*., 2018; Bunketorp-Käll *et al*., 2019, 2020) fulfilled at least four out of the five quality criteria. However, other studies were lacking in major methodological details, suggesting that they had not been met. Major risks of bias include inappropriate randomization or insufficient information regarding randomization in five out of eight RCT. Other concerns include missing data, blinding and incomplete information about adherence to assigned intervention.

**Tab. 2.**
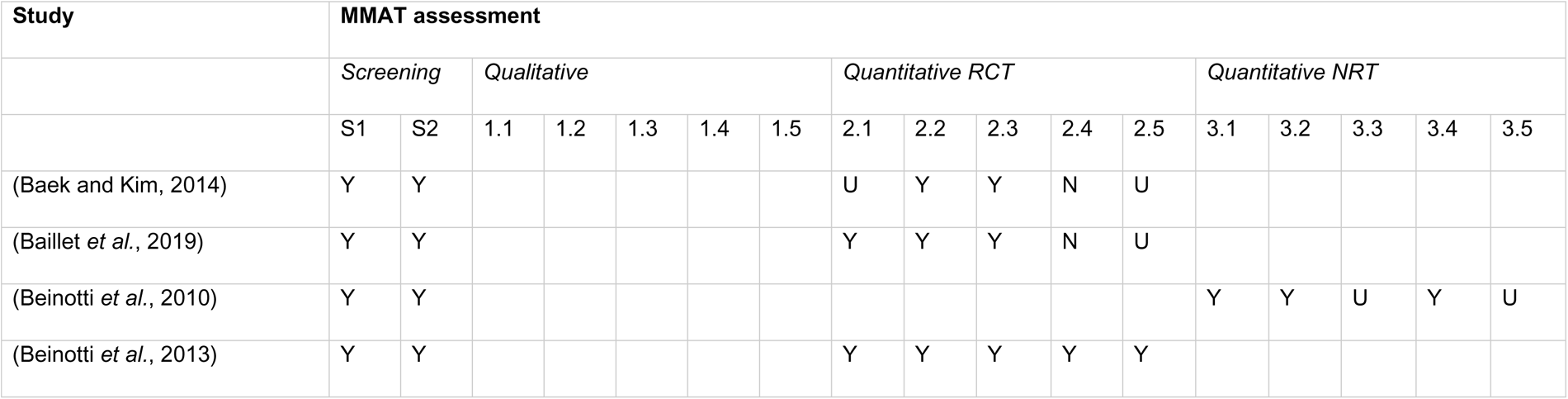
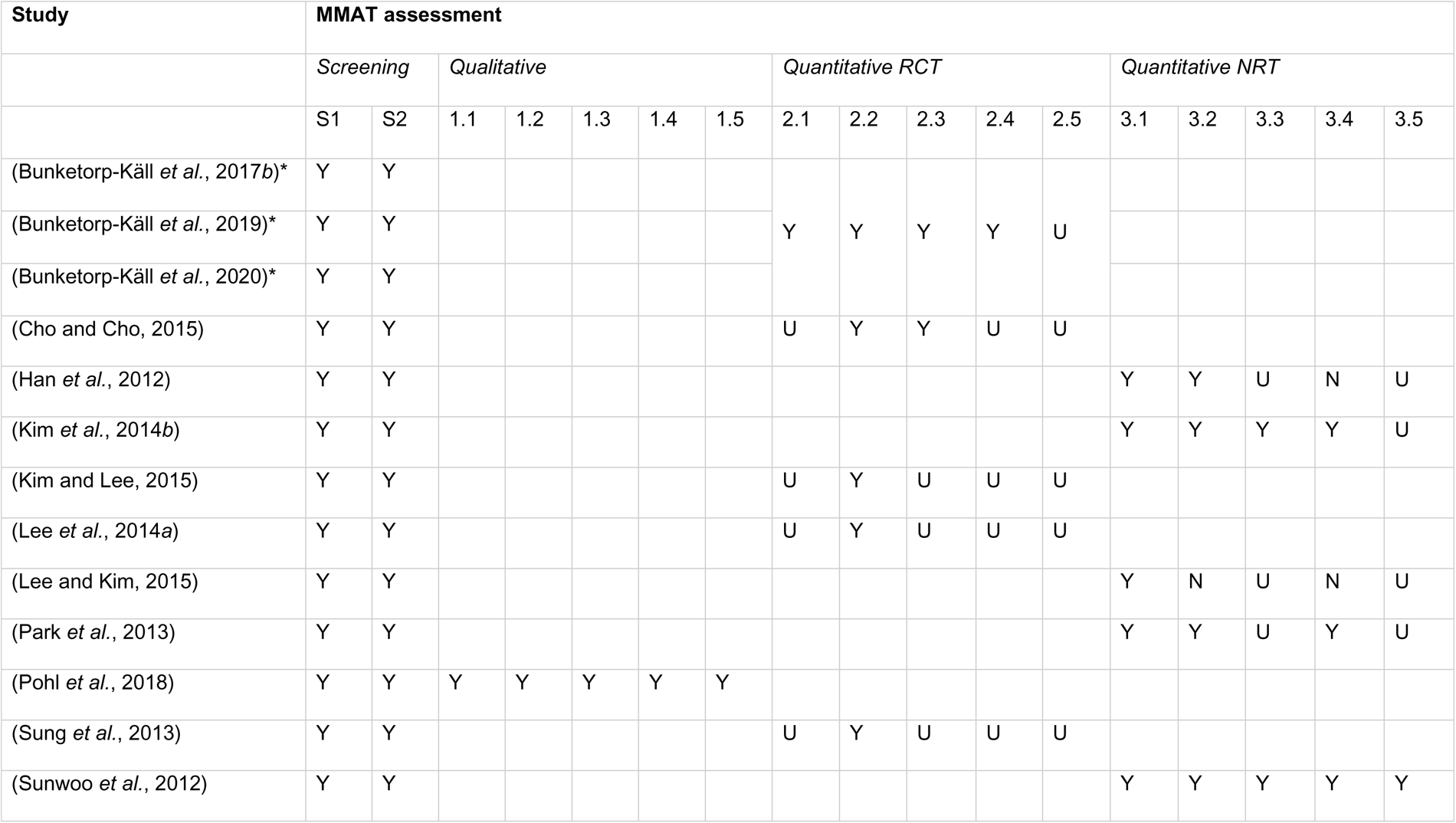
Overview of study quality. Study quality was assessed by MMAT. Depending on the method of the study, quality was assessed using different questions. Questions: S1: Are there clear research questions?, S2: Do the collected data allow to address the research questions?, 1.1: Is the qualitative approach appropriate to answer the research question?, 1.2: Are the qualitative data collection methods adequate to address the research question?, 1.3: Are the findings adequately derived from the data?, 1.4: Is the interpretation of results sufficiently substantiated by data?, 1.5: Is there coherence between qualitative data sources, collection, analysis and interpretation?, 2.1: Is randomization appropriately performed?, 2.2: Are the groups comparable at baseline?, 2.3: Are there complete outcome data? [at least 80% of data], 2.4: Are outcome assessors blinded to the intervention provided?, 2.5: Did the participants adhere to the assigned intervention?, 3.1: Are the participants representative of the target population?, 3.2: Are measurements appropriate regarding both the outcome and intervention (or exposure)?, 3.3: Are there complete outcome data?, 3.4: Are the confounders accounted for in the design and analysis?, 3.5: During the study period, is the intervention administered (or exposure occurred) as intended? (Hong *et al*., 2018) Abbreviations: MMAT = Mixed Methods Appraisal Tool, N = criterion not met, NRT = Non-randomized Trial, RCT = Randomized Controlled Trial, U = unclear if criterion was met, Y = criterion met *Findings from data of same study.

### Intervention

Different forms of intervention and terminology were used. Four studies referred to their intervention as *hippotherapy* (Beinotti *et al*., 2010; Sunwoo *et al*., 2012, Lee *et al*., 2014*b*) or *hippotherapy simulator* (Sung *et al*., 2013); four studies used *horse-riding therapy* (Bunketorp-Käll *et al*., 2017*b*, 2019, 2020) or *horse-riding exercise* (Kim and Lee, 2015); two studies used *horseback riding* (Pohl *et al*., 2018) or *horseback riding therapy* (Beinotti *et al*., 2013); two studies used *horse riding simulation training* (Baek and Kim, 2014, Kim *et al*., 2014*a*); one study used *horseback riding simulator exercise* (Park *et al*., 2013), a further study used *mechanical horse practice* (Baillet *et al*., 2019); and three studies referred to their intervention as *mechanical horseback riding* (Han *et al*., 2012; Cho and Cho, 2015; Lee and Kim, 2015). The exact protocol was not, or only briefly described in most studies. In general, some studies included exercises during riding while other studies did not.

A short summary of the parameters of EG and CG applied in each study can be found in Tab. 1. While eight studies applied EAT in addition to conventional therapy (Beinotti *et al*., 2010; Han *et al*., 2012; Beinotti *et al*., 2013; Park *et al*., 2013; Sung *et al*., 2013; Baek and Kim, 2014; Cho and Cho, 2015; Lee and Kim, 2015), the others applied EAT only. (Sunwoo *et al*., 2012, Kim *et al*., 2014*a*, Lee *et al*., 2014*a*; Kim and Lee, 2015, Bunketorp-Käll *et al*., 2017*b*; Pohl *et al*., 2018; Baillet *et al*., 2019; Bunketorp-Käll *et al*., 2019, 2020) Total time of EAT varied between 300 and 5760min (1043 ± 1377min, Mean ± SD). EAT was applied within four to 16 weeks, in 16 to 30 total sessions.

With the exception of two studies (Sunwoo *et al*., 2012, Kim *et al*., 2014*a*), all included a CG which consisted of treadmill training (Lee *et al*., 2014*a*) or another form of conventional therapy in the other studies. While most of the studies had comparable doses for the EG and CG, in four studies, both groups performed the same training and the EG did EAT in addition; thus resulting in different training doses. (Han *et al*., 2012; Beinotti *et al*., 2013; Cho and Cho, 2015; Lee and Kim, 2015)

In five studies, the intervention was applied with horses (Beinotti *et al*., 2010; Sunwoo *et al*., 2012; Beinotti *et al*., 2013, Lee *et al*., 2014*a*, Bunketorp-Käll *et al*., 2017*b*; Pohl *et al*., 2018; Bunketorp-Käll *et al*., 2019, 2020), whereas the other studies used riding simulators.

### Outcome measures

#### Physical measures

Thirteen studies evaluated the effects of EAT on physical outcome measures.^14,40,42– 52,55^ The domains evaluated included balance, gait, postural coordination, activities of daily living (ADL), lower extremity motor impairment, motor function and hand strength.

Balance was measured in eleven studies.^8,14,42,45–52^ It was assessed by the Berg Balance Scale (BBS), Balance Part of the Performance Oriented Mobility Assessment (B-POMA, POMA), Balance systems, Balance Items of the Fugl-Meyer Lower Extremity Scale (FMLE), and Bäckstrand, Dahlberg and Liljenäs Balance Scale (BDL-BS) and the Trunk Impairment Scale (TIS).

Gait was assessed in nine studies.^8,40,42,43,45–47,49–51^ It was investigated by the Functional Ambulation Category Scale (FAC), Functional Gait Assessments (FGA; part cadence), Timed Up and Go Test (TUGT), Timed 10-meter walk test (10 mWT), the six-minute walk test (6 MWT), Gait part of Performance Oriented Mobility Assessment (G-POMA), and Gait analyzer (measures: Cadence, Double Support, Load Response, Pre-Swing, Single Support, Stance Phase, Step Length Asymmetry, Step Length, Stride, Swing Phase, Velocity).

In one study, an optical tracking system was used to assess postural coordination.^44^ ADL were investigated in two studies using the Modified Barthel Index (MBI).^45,50^

In one study, general motor function was investigated using the Modified Motor Assessment Scale according to Uppsala University Hospital (M-MAS UAS) and hand strength was assessed using a dynamometer (Grippit)^8,40^ In another study, lower extremity motor impairment was investigated by the FMLE.^46^

#### Further health-related measures

The effects on further health-related outcome measures were investigated in seven studies.^8,14,18,41,43,45,51,53^ The measures included depression, perception of the intervention and of recovery from stroke, cognition, quality of life (QoL), muscle thickness, as well as trunk muscle activity.

Depression was assessed in two studies by Becks Depression Inventory (BDI) and/or Hamilton Depression Rating Scale (HAM).^45,51^ Another study assessed different domains of the patients’ perception of EAT. Specifically, a qualitative assessment using semi-structured interviews was performed in one report^53^, and the correlation between changes in different outcomes and the change in perceived recovery from stroke was estimated in another report.^41,53^ Furthermore, the change in perceived recovery from stroke (Stroke Impact Scale, Item 9; SIS-9) after EAT was also assessed in this study. Moreover, both the general cognitive level and the working memory were examined using the Barrow Neurological Institute screen for higher cerebral functions and the Letter-number sequencing test, respectively.^8^ QoL was investigated in one study by the Medical Outcomes Study 36-item Short-Form health survey (SF-16).^18^

One study examined muscle thickness using ultrasonic imaging.^14^ The other study investigated trunk muscle activity by electromyography (EMG).^43^

### Effects of intervention on outcome measures

#### Effects of EAT on physical outcome

Our search revealed studies identifying effects of EAT on balance and gait. Therefore, an overview of within- and between-effects of EAT on both balance and gait is shown in Fig. 2 and Fig. 3, respectively. While different study designs (RCT and NRT) are indicated in these plots, results appeared to be independent of the study design. Therefore, in the following, findings are described for all included studies together.

**Fig. 2.**
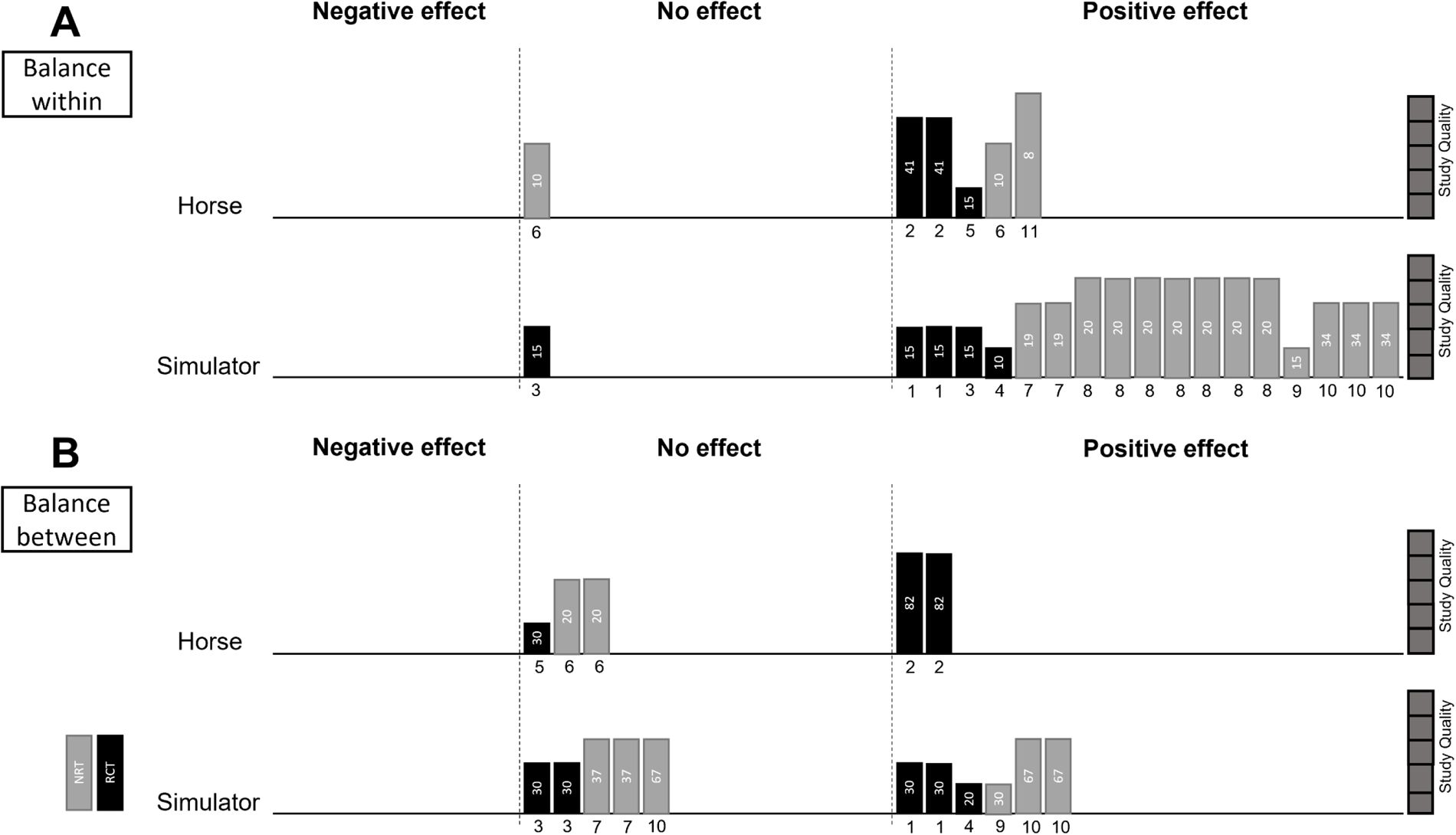
Harvest Plot: Effects of EAT on balance. A) shows within-effects of EAT on balance, and B) shows between-effects. Both in A) and B) each bar represents one outcome related to balance of one study (in case one study assessed balance using multiple outcome measures, each is represented in a separate bar; the study identification is indicated by a number below the bar). In the upper row, studies applying EAT with horses are shown, and in the lower row, studies applying EAT with simulators are show. The colour of the bars indicates the study design (RCT: black, NRT: grey). The height of the bar represents the study quality, assessed by the Mixed Methods Appraisal Tool. The number in the box represents the sample size. Studies are grouped according to a negative (left), no (middle), or a positive effect (right), based on the reported statistical test results in the respective papers. Studies: 1: Baek and Kim 2014, 2: Bunketorp-Käll *et al*. 2017, 3: Cho and Cho 2015, 4: Kim and Lee 2015, 5: Lee *et al*. 2014, 6: Beinotti *et al*. 2010, 7: Han *et al*. 2012, 8: Kim *et al*. 2014, 9: Lee and Kim 2015, 10: Park *et al*. 2013, 11: Sunwoo *et al*. 2012

**Fig. 3.**
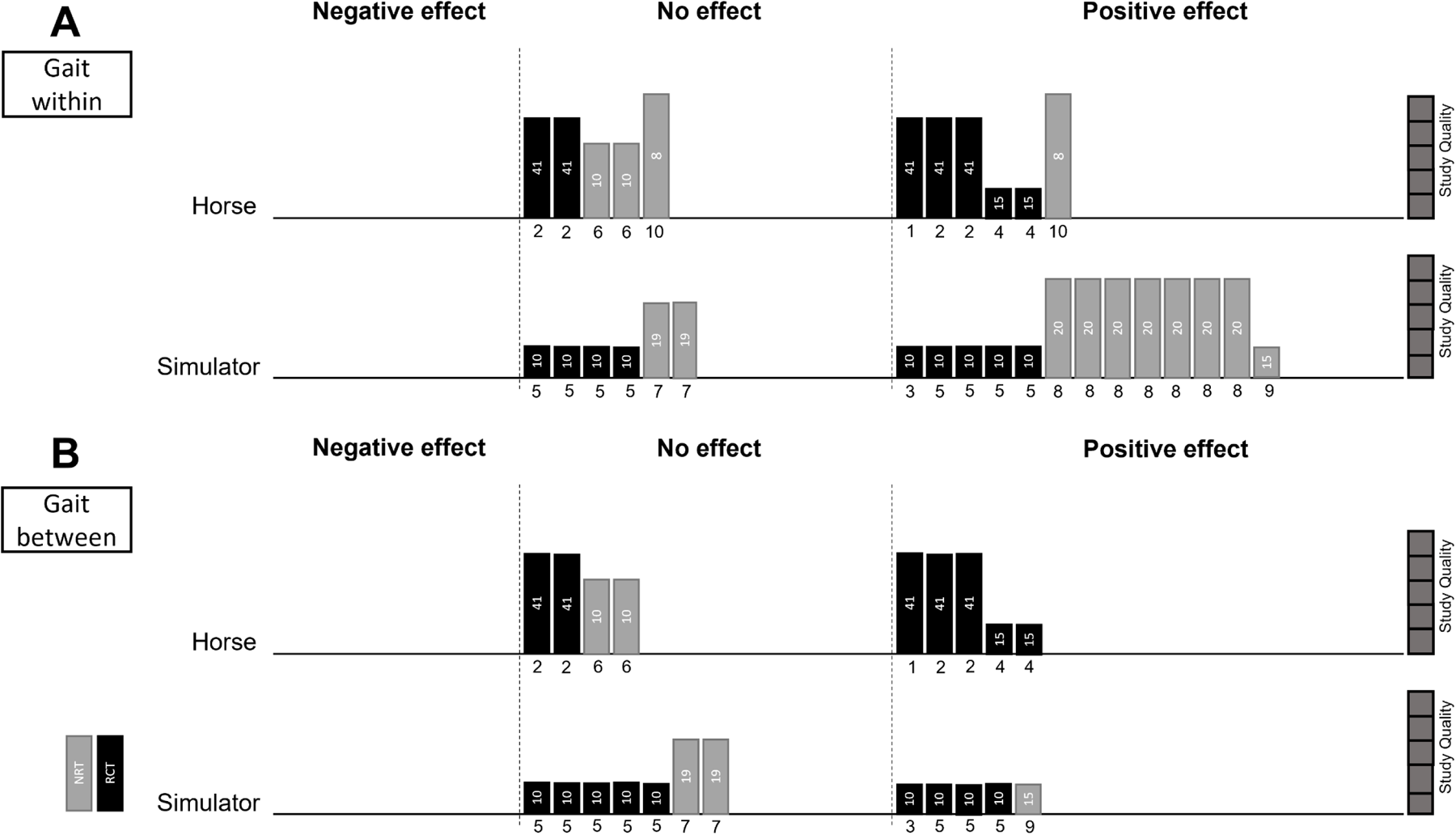
Harvest Plot: Effects of EAT on gait. A) shows within-effects of EAT on gait, and B) shows between-effects. Both in A) and B) each bar represents one outcome related to gait of one study (in case one study assessed gait using multiple outcome measures, each is represented in a separate bar; the study identification is indicated by a number below the bar). In the upper row, studies applying EAT with horses are shown, and in the lower row, studies applying EAT with simulator are show. The colour of the bars indicates the study design (RCT: black, NRT: grey). The height of the bar represents the study quality, assessed by the Mixed Methods Appraisal Tool. The number in the box represents the sample size. Studies are grouped according to showing a negative (left), no (middle), or a positive effect (right), based on the reported statistical test results in the respective papers. Studies: 1: Bunketorp-Käll *et al*. 2017, 2: Bunketorp-Käll *et al*. 2019, 3: Kim and Lee 2015, 4: Lee *et al*. 2014, 5: Sung *et al*. 2013, 6: Beinotti *et al*. 2010, 7: Han *et al*. 2012, 8: Kim *et al*. 2014, 9: Lee and Kim 2015, 10: Sunwoo *et al*. 2012

Balance was usually assessed by BBS, i.e., in eight out of eleven studies. All of these reported significant within-effects (Cohen’s d = 0.22 to 2.23) (Beinotti *et al*., 2010; Han *et al*., 2012; Sunwoo *et al*., 2012; Park *et al*., 2013, Lee *et al*., 2014*a*; Kim and Lee, 2015; Lee and Kim, 2015, Bunketorp-Käll *et al*., 2017*b*). Moreover, significant within-effects in POMA (Sunwoo *et al*., 2012) (Cohen’s d = 0.44), B-POMA (Han *et al*., 2012) (Cohen’s d = 0.93), BDL-BS (Cohen’s d = 0.89) (Bunketorp-Käll *et al*., 2017*b*) and TIS (Kim *et al*., 2014*a*) (Cohen’s d = 1.43) were observed after EAT. With balance systems, significant effects on balance could be identified, specifically on Center of Pressure (COP) path length (Cohen’s d = 0.78) and COP travel speed (Cohen’s d = 0.39) (Baek and Kim, 2014), EC moving distance of COP (Cohen’s d = 0.63) (Cho and Cho, 2015), EO and EC sway length (Cohen’s d = 0.67 and 1.13, respectively), EO and EC sway speed (Cohen’s d = 0.46 and 1.13, respectively) (Kim *et al*., 2014*a*), and EO and EC sway area (Cohen’s d = 0.35 and 0.50, respectively) (Kim *et al*., 2014*a*); EO balance (Cohen’s d = 0.08) and EC balance (Cohen’s d = 0.71) (Park *et al*., 2013). However, no significant effect was found for EO moving distance of COP (Cohen’s d = 0.33) (Cho and Cho, 2015) or for the balance part of FMLE (Cohen’s d = 0.14) (Beinotti *et al*., 2010).

Moreover, nine studies assessed between-effects of EAT on balance. Improvements in BBS were found to be significantly higher in EG than CG in four out of seven studies (Cohen’s d between 0.57 and 1.66) investigating between-effects on BBS. (Park *et al*., 2013; Kim and Lee, 2015; Lee and Kim, 2015, Bunketorp-Käll *et al*., 2017*b*) In addition, significant between-effects on balance were found for COP Path Length and Travel Speed (Cohen’s d = 0.68 and 0.39, respectively) (Baek and Kim, 2014), the BDL-BS (Cohen’s d = 0.11) (Bunketorp-Käll *et al*., 2017*b*), EC balance (Cohen’s d = 0.31) (Park *et al*., 2013). However, no significant between-effects were found for FMLE-balance (Cohen’s d = 0.07) (Beinotti *et al*., 2010), EO and EC COP Moving Distance (Cohen’s d = 0.01 and 0.31, respectively) (Cho and Cho, 2015), B-POMA (0.95) (Han *et al*., 2012), or EO balance (Cohen’s d = -0.04) (Park *et al*., 2013).

For gait, positive within-effects of EAT were noted in the TUGT (Cohen’s d between 0.48 and 1.84) (Lee and Kim, 2015, Bunketorp-Käll *et al*., 2017*b*) and the 10 mWT (Cohen’s d between 0.26 and 1.55) (Sunwoo *et al*., 2012; Kim and Lee, 2015; Bunketorp-Käll *et al*., 2019). Moreover, with gait analyzer, positive effects were found on velocity (Cohen’s d = 0.59 – 2.06) (Kim *et al*., 2014*a*, Lee *et al*., 2014*a*), stride of the affected and unaffected sides (Cohen’s d = 0.35 and 0.29, respectively), Double Support of the affected and unaffected sides (Cohen’s d = 0.59 and 0.31, respectively) (Kim *et al*., 2014*a*), Step Length Asymmetry (Cohen’s d = 2.60) (Lee *et al*., 2014*a*), single support (Cohen’s d = 2.34), load response (Cohen’s d = 1.61), total double support (Cohen’s d = 1.07), and pre-swing (Cohen’s d = 1.76) (Sung *et al*., 2013). However, no effects of EAT on gait were detected in the 6 MWT (Cohen’s d = 0.68) (Bunketorp-Käll *et al*., 2019), the FAC (Cohen’s d between 0.00 and 0.23) (Beinotti *et al*., 2010; Han *et al*., 2012; Sunwoo *et al*., 2012), or on the G-POMA (Cohen’s d = 0.00) (Han *et al*., 2012). In addition, with gait analyzer, no effects of EAT were found on step length (Cohen’s d = -0.01), stance phase (Cohen’s d = 0.13), or swing phase (Cohen’s d = 0.49) (Sung *et al*., 2013). For cadence, one study found a positive effect (Kim *et al*., 2014*a*) (Cohen’s d = 0.33; assessed by gait analyzer), while others reported a negative effect (Beinotti *et al*., 2010; Sung *et al*., 2013) (Cohen’s d = -0.16, assessed by FGA; and Cohen’s d = 0.00, gait analyzer).

Moreover, between-effects of EAT on gait were investigated for the following gait parameters: FAC, Cadence, TUGT, 10 mWT, 6 MWT, G-POMA as well as on gait velocity, step length asymmetry, step length, stance phase, swing phase, load response, single support, total double support, pre-swing and cadence using Gait Analyzer. Significant between-effects were found for the 10 mWT (Cohen’s d = 0.08 – 1.31) (Kim and Lee, 2015; Bunketorp-Käll *et al*., 2019), TUGT (Cohen’s d = 1.85) (Lee and Kim, 2015), Gait velocity (Cohen’s d = 1.42), Step Length Asymmetry (Cohen’s d = 1.96) (Lee *et al*., 2014*a*), Load Response (Cohen’s d = 0.64), Single Support (Cohen’s d = 1.86), and Pre-Swing (Cohen’s d = 0.86) (Sung *et al*., 2013).

In addition, benefits of EAT on posture were found within a pilot study (Cohen’s d 1.04 – 3.74). (Baillet *et al*., 2019)

Of the two studies investigating changes in ADL after EAT, one study detected a significant improvement in ADL (Cohen’s d = 1.96) (Kim and Lee, 2015), while the other study did not find an effect on ADL (Cohen’s d = 0.12) (Sunwoo *et al*., 2012).

In one study, lower extremity motor impairment improved significantly in the EG (Cohen’s d = 1.03), and these improvements were also significantly larger than for CG (Cohen’s d = 0.66). (Beinotti *et al*., 2010)

Moreover, there was a significant improvement in overall motor function after EAT in one study, when comparing EG and CG (Cohen’s d = 0.23). (Bunketorp-Käll *et al*., 2019)

However, there was no significant effect of EAT on hand strength. (Bunketorp-Käll *et al*., 2017*b*)

#### Effects of EAT on further health-related outcome

Findings for depression varied between studies. One study reported significant improvements in the BDI after EAT^51^, for the comparisons pre to post EAT, and EG to CG (Cohen’s d = 1.58 and 1.25, respectively), while the other did not show any significant changes for the BDI or the HAM.^45^

Two studies investigated the perception of the intervention itself, which was reported to have positive effects on different domains.^53^ To summarize, stroke patients perceived the intervention as a rich and pleasurable experience that had a positive impact on their emotional and physical domains.^53^ Furthermore, changes in perceived recovery were associated with improved aspects of gait.^41^

Perceived recovery from stroke increased significantly after EAT and persisted for three- and six-months post intervention (Cohen’s d = 0.78, 3-months post intervention). This increase was furthermore significantly higher than for the CG (Cohen’s d = 0.95).^8^

Although there were slight increases in general cognition and the Letter Number Sequencing test in one study, no significant effect was observed for these parameters after EAT (Cohen’s d = 0.30 and 0.17, respectively).^8^

There was a positive effect on QoL after EAT, in particular for the domains of functional capacity, physical aspects and mental health.^18^

Prior to EAT, the external oblique muscle on the affected side was significantly less thick than on the non-affected side, as measured by ultrasonic imaging. Following EAT, the thickness increased significantly (Cohen’s d = 0.33), and was therefore more similar to the non-affected side. By contrast, muscle thickness of the internal oblique and transversus abdominis was comparable between affected and non-affected side and did not change after the intervention.^14^

In addition, by using EMG recordings, the trunk muscle activity during sit-to-stand was measured after EAT. Paretic erector spinae activation was significantly higher for the EG after the intervention (Cohen’s d = 0.91) which, moreover, differed significantly from the CG (Cohen’s d = 1.20). By contrast, the rectus abdominis remained unchanged.^43^

#### Difference in physical outcome when applying EAT with horses or riding simulators

In general, we could only identify five studies applying EAT after stroke with horses. It was only possible to compare the differences for balance and gait.

For within-effects on balance, four studies applied EAT with horses and seven studies in simulators (see Fig. 2). Positive within-effects were found for most studies/ outcomes, independent of EAT with horses or simulators, study quality or design. Three vs. six studies investigated between-effects on balance with horses and simulators, respectively. Significant between-effects on balance were found for one RCT with high methodological quality and a large sample size applying EAT with horses but not in two other studies (one RCT and one NRT) with lower methodological quality and small sample sizes. For studies applying EAT with riding simulators, results varied. However, in addition to the unequal number of reports to compare, studies were highly heterogeneous in terms of study design, quality, and sample size.

Effects of EAT on gait were evaluated in five reports with horses and in five reports with riding simulators (see Fig. 4). Four studies each investigated also between-effects. Independent of the study design, for both horses and simulators, significant and non-significant effects on gait were detected. However, studies were highly heterogeneous similarly to the studies on balance. Furthermore, outcome measures were inconsistent between studies.

## Discussion

### Summary of results

Taken together, this systematic literature review revealed 14 studies that evaluated the effect of EAT on health-related outcomes. While thirteen studies reported physical outcomes, seven studies reported further health-related outcomes. In general, the positive effects of EAT on stroke recovery could be identified in all the domains investigated, i.e., physical (balance, gait, postural control, lower extremity motor impairment, motor function, and ADL) and further outcomes (perceived recovery from stroke, and QoL, abdominal muscle thickness, trunk muscle activation). The most consistent and robust beneficial effects were identified on balance and gait.

### Physical effects of EAT

Although different aspects of physical effects of EAT were studied, most of the studies focused on investigating the effects of EAT on balance and/or gait.

Independent of the exact intervention, dose and study design, all eight studies investigating within-effects of EAT on balance, as measured by BBS, reported positive effects of EAT. Furthermore, in four out of seven studies assessing between-effects of EAT on balance (measured by BBS), significant improvements were reported.^8,50– 52^ BBS is a valid tool for assessing balance during stroke recovery with an excellent reliability.^56^ It measures both the static and dynamic aspects of balance, i.e., to maintain balance either statically or during functional tasks.^56^ The 14 items include tasks such as standing on one or two legs with eyes open and closed, sitting or standing up or sitting down.^57^ This suggests a robust effect of EAT on general balance which has previously also been supported by a meta-analysis (albeit only including two studies).^58^ In addition, positive findings were described for individual aspects of balance, which were assessed by balance systems. However, these studies investigated different aspects of balance and were in general of low methodological quality. To conclude, EAT appears to have a positive effect on general balance, whereas the exact details require further consideration.

After stroke, gait issues are very common. Specifically, reduced walking speed and longer stance phase have been reported after stroke.^59,60^ After EAT, walking speed (measured by different assessment tools) was consistently found to increase.^8,40,42,45,49–51^ Moreover, these improvements were higher in the EG than in the CG in some studies.^8,40,42,50,51^ This indicates a robust finding across different studies, independent of study design and quality. However, for other gait parameters, especially those investigated by Gait Analyzer, evidence is sparse, and conclusions are restricted due to limitations in study quality.

Taken together, there is considerable evidence for a positive effect of EAT on balance and gait. Recovery of gait parameters was reported as a major goal of stroke patients and is associated with quality of life.^61^ In addition, recovery of gait and balance may decrease the risk of falling in stroke patients.^60,62^ These findings support the promising potential of EAT as a multimodal intervention for recovery after stroke. In addition, preliminary evidence was provided for improved lower extremity motor impairment^46^ and overall motor function after EAT^40^. However, to our knowledge, the effects of EAT on fine motor control or spasticity in stroke patients have yet to be evaluated. While fine motor control was not formally assessed, stroke patients reported subjectively improved fine motor skills during a semi-structured interview.^53^ Moreover, temporary (lower extremity) spasticity was shown to decrease for patients with cerebral palsy^10,63^ or spinal cord injury^13,25^. Even though the effects on upper extremity spasticity remain unclear, EAT may hold a promising potential for decreasing spasticity after stroke. Future research should investigate both short- and long-term changes of upper and lower extremity spasticity, as well as changes in fine motor control.^21,30^

### Effects of EAT on further health-related outcomes

Different further effects after EAT were investigated in stroke patients. However, evidence was sparse, and findings varied between studies. For example, EAT was found to have positive effects on perceived recovery from stroke and QoL.^8,18^ Furthermore, the intervention (applied in horses) had a large emotional impact on the participants. For example, they reported higher self-esteem and increased self-efficacy.^53^ Moreover, perceived recovery from stroke was associated with improved gait parameters, emphasizing the high subjective importance of gait recovery for stroke patients.^41^

The evidence for effects of EAT on depression was, however, inconclusive^45,51^ While depression was shown to improve in one study^64^, it did not change on the other study^45^. In this pilot study, however, the dose of EAT was much lower than in the other studies (i.e., 8h of EAT^45^ vs. 15h EAT^64^) Additionally, only eight patients were enrolled in this study. Moreover, the study that showed significant improvements of depression was of low methodological quality. The results might therefore be compromised by these constraints and require further consideration. In addition, no significant effects of EAT on cognitive aspects were observed. However, cognitive outcome was assessed in only one study.^8^

Moreover, it is important to understand the underlying physiological mechanisms of EAT to explain the findings and to predict and optimize EAT protocols. In this context, our systematic literature search revealed only two studies for stroke patients; these studies investigate muscle thickness and activity.^14,43^ It was found that muscle activity and thickness of the paretic side change after EAT which supports theories that EAT is training respective muscles.^17,18^ However, it should be noted that study quality was low in both studies.

While these studies may help to explain physical changes, e.g., in gait or balance, other factors may shape our understanding of the mechanisms of action and should be investigated in future studies. In healthy, elderly people, for example, EAT led to hormonal changes, i.e., to a significant increase in serotonin and a decrease in cortisol levels.^65^ In an autistic population, a decrease in the level of cortisol was detected in the course of EAT training.^66^ In addition, an increase of progesterone levels over time was found.^66^ Cortisol is a classical biomarker of stress and may thus reflect a lower level of stress after EAT.^67^

Moreover, preliminary investigations of brain physiology in this context have been made using EEG, fMRI, and NIRS.^27,68–71^ In healthy elderly people, alpha power in the EEG increased during both riding a horse^69^, and a riding simulator.^69,70^ Furthermore, one study used fMRI to assess changes in brain connectivity in children with ADHD.^27^ Increased connectivity was detected both within the cerebellum (albeit not statistically higher than in the CG)^72^ and from the cerebellum to other regions, i.e., to the right insular cortex, right middle temporal gyrus, left superior temporal gyrus, and right precentral gyrus^27^.

In addition, heart rate variability (HRV) in older adults increased after EAT when interacting with horses.^73^ Interestingly, the subjects’ HRV often synchronized with the horses’ HRV, suggesting social bonding.^73,74^

Taken together, these findings suggest a positive effect of EAT on relaxation and increased concentration.^67,69,73,74^ However, it remains unclear as to which specific brain areas and networks are active and affected by EAT in stroke patients. Understanding the physiological mechanisms and clinical correlates of EAT may help to further optimize the therapy as well as to stratify patients who may benefit from it.^75^

### EAT applied with horses vs. riding simulator

To assess the whole field, we included studies that applied EAT in both riding simulators and horses. These approaches have different advantages and disadvantages:

Although similar, riding simulators may not reflect movements comparable to those of a horse.^76^ For example, some simulators apply only two-dimensional movements.^44,76^ On the other hand, simulators offer the advantage of rhythmic movements without any deviation from the protocol compared to training with a real animal.^44^ Riding simulators may also be more easily accessible and more affordable.^76^ However, the emotional and psychological effects of the interaction with a horse should not be underestimated. Meaningful and positively stimulating training may increase the likelihood of improvement.^53,77,78^ For example, stroke patients who underwent EAT for several weeks stressed the importance and strong effect of bonding with the horse during therapy.^53^ This was also confirmed by HRV studies which found a favorable effect of EAT on HRV.^67,73,74^

Both horses and riding simulators seem to yield positive effects on balance and gait without favoring one over the other. Consequently, based on previous literature on equine-assisted therapy (EAT) after stroke, it is challenging to determine the superiority of one approach over the other. Additionally, investigating their impact on other factors would be of significant interest and should be explored in future studies.

Prior research suggests that the benefits of applying EAT in horses may outweigh those of riding simulators.^44,76,79^ However, for introducing patients to the movements and training, using riding simulators could prove beneficial.

### Study Quality

The study quality assessment suggests low methodological quality for most studies. Risk of bias include inappropriate randomization in RCT, missing data, blinding and incomplete information about adherence to assigned intervention. Therefore, findings from previous research as well as the presented synthesis of the results cannot be generalized to all stroke patients and require further investigation.

However, although most current evidence is limited by methodological constraints, seven reports (based on five studies) were of considerable methodological quality. Four of them reported improved gait^8,40,45,49^ and three of them improved balance^8,45,49^ after EAT. Similar effects have been identified for EAT in children with cerebral palsy^22,80^ which may have similarities to stroke patients. Therefore, synthesized results of this review, especially the effectiveness of EAT in boosting gait and balance after stroke, should not be disregarded but rather taken as a starting point for further investigation.

### Limitations

We are aware of multiple limitations regarding this systematic review. In general, previous evidence for the effects of EAT in stroke patients is sparse.

To comprehensively assess the entire field, our goal was to incorporate all studies related to Equine-Assisted Therapy (EAT) during stroke recovery. The term EAT encompasses various specific therapies involving horses, ^13,17^ such as those focusing on teaching riding skills, improving motor function, or providing psychotherapy. ^13^

Importantly, inconsistent use of terms and often insufficiently detailed descriptions of interventions make it challenging to distinguish between their respective effects. Additionally, different interventions may exhibit variations in their effects. ^13,17^ We also included studies with diverse study designs.

Consequently, the included studies exhibited high heterogeneity in terms of study design, study quality, applied intervention (horses or simulators), intervention dosage, and control groups (see Table 1, Fig. 3 and 4). Due to this substantial heterogeneity, we were unable to draw overarching conclusions. To address this diversity, our approach was to synthesize the literature based on study design and whether the studies employed EAT with horses or riding simulators.

## Conclusion and future directions

To conclude, EAT combines several important factors that may boost stroke rehabilitation of different symptoms. Specifically, this multimodal intervention consists of sensory, motor and cognitive components, offers high intensity training, and incorporates emotional and motivational aspects.^8,12^

While the benefits of EAT on recovery after stroke were identified in different domains, previous research is sparse and methodologically limited. This highlights the need of future research which should especially focus on systematically evaluating the effects of EAT on physical function including upper extremity motor function and spasticity, as well as on psychological factors. In addition, investigating physiological changes may help us to gain a better understanding of the underlying mechanisms and to further optimize future therapies.

## Authorship contribution statement

BHT: Conceptualization, Methodology, Investigation, Formal Analysis, Visualization, Writing – Original Draft, AG: Conceptualization, Project administration, Funding acquisition, Writing – Review and Editing

## Acknowledgments

This work was supported by the German Federal Ministry of Education and Research [BMBF 13GW0570, BEVARES]. We acknowledge support from the Open Access Publishing Fund of the University of Tuebingen. We thank Karolina Talar for valuable methodological input.

## Data availability

The data that support the findings of this study are available for researchers from the first author upon reasonable request.

